# Assessing the Impact of a NICU Book Cart on Staff and Caregiver Speech to Very Preterm Infants

**DOI:** 10.1101/2025.11.26.25341053

**Authors:** Sasha Montgomery, Avery Look, Molly Lazarus, Virginia A. Marchman, Melissa Scala, Katherine E. Travis

## Abstract

**Objective:** To examine differences in infant-directed talk activities (e.g., reading) before and after introduction of a NICU book cart.

**Study Design:** We retrospectively analyzed medical record data of infants born <32 weeks gestational age (*n* = 147) whose NICU stay occurred in the year before or after book cart implementation (6/1/2022). Talk rates and frequencies were calculated from clinically charted family and staff talk data; talk amounts were compared across pre- and post-book cart cohorts, adjusting for covariates.

**Results:** Cohorts displayed equivalent clinical/demographic variables, except for higher bronchopulmonary dysplasia rates in the post-book cart cohort. Post-book cart family and staff engaged in more talk than those in the pre-book cart cohort, with staff showing higher talk rates and frequencies than family across both cohorts.

**Conclusion:** Family and staff with access to a NICU book cart engaged in more talk, suggesting a strategy to support infant speech exposure.

## Introduction

Preterm birth is associated with increased rates of language impairments that can persist throughout the lifespan.^1–3^ Compared to children born at term, children born preterm often demonstrate weaker early language skills, including delayed lexical and grammatical development that, in turn, can negatively impact later academic abilities such as reading.^1,4^ Speech perceptual abilities, considered precursors to later language skills, first emerge in the late second and third trimesters and are thought to be shaped by regular exposure to maternal speech sounds in the womb.^5,6^ For infants born preterm, these early auditory experiences are often substantially reduced during prolonged hospitalizations in the neonatal intensive care unit (NICU)^7^. Reduced speech exposure may therefore contribute to alterations in brain development and language delays observed in preterm children.^8^ Identifying interventions that increase speech exposure during the NICU period is needed if we are to support early brain development and language outcomes in these vulnerable children.

A common strategy to increase language exposure and support early language development in infants and toddlers is shared reading.^9–13^ Clinicians are often trained to encourage parents to read to their infants during outpatient visits, and there is evidence that such practices can positively influence language outcomes as early as 9 months of age.^14^ Programs like *Reach Out and Read* have demonstrated that providing books and promoting shared reading not only enhances parental literacy behaviors but also improves children’s language skills.^15–17^ In the NICU setting, experimental studies have shown that shared reading with preterm infants can foster parent-infant bonding, reduce parental stress, and promote language development.^18–23^ Many hospitals now partner with nonprofit organizations to distribute donated books to families in the NICU. Yet, the impact of these initiatives on the overall speech environment remains unclear. Specifically, it is unknown whether providing books outside of structured research interventions increases the amount of speech families provide to their infants. It is also unclear whether access to books influences the verbal behaviors of clinical staff, who may be key contributors to the infant’s auditory environment due to their consistent bedside presence.

Understanding whether increasing book access with minimal intervention can enrich the NICU speech environment is important for informing low-cost, scalable strategies to support brain and language development in at-risk infants.

To address these questions, the present study evaluated whether the presence of a book cart in the NICU influenced how often and how much families and clinical staff engaged in talk activities. We hypothesized that family and clinical staff would talk more in the period after the book cart became available than in an equivalent period prior to the placement of the book cart. Based on our previous findings that clinical staff produce higher speech counts during care times than family members,^24^ we also anticipated that clinical staff would contribute more speech overall than families.

## Methods

### Study Design

We performed a retrospective study, analyzing electronic medical record (EMR) data collected as part of routine clinical care in our unit. This study was considered a minimal-risk retrospective chart review; families were not required to provide consent (Stanford University IRB #44480).

### Sample

Participants were infants (*N*=147; 68 females, 79 males) born very preterm (< 32 weeks GA) who were cared for at Lucile Packard Children’s Hospital in a 72-bed level IV NICU with an open bay design. Data were obtained from an existing clinical database that contained all very preterm infants who had undergone routine brain MRI scanning close to the time of hospital discharge. From this database, infants were selected to be in the current study if they were admitted to our center within the first 7 days of life, had no congenital anomalies, and were discharged from our center at a term equivalent age. Even though the book cart was stocked with some picture books, the worded books were in either English or Spanish. Therefore, we excluded infants (n=9) if their family’s primary language did not include Spanish or English. Infants were categorized into one of two cohorts; (1) the pre-book cart group (n=66) included infants who completed their entire NICU hospitalization in the year immediately prior to book cart installation (5/1/2021 and 5/1/2022); (2) the post-book cart group (n=81) included infants born and discharged in the year after the book cart was installed and fully stocked (6/1/2022-6/1/2023).

## Procedure and Measures

### Book Cart Installation

A wooden book cart containing children’s picture books or word books in English or Spanish was installed in the unit starting 5/1/2022 and was fully stocked with books by 6/1/2022. Both clinical staff and families were informed of the addition of the book cart to the NICU. Books were freely available for families to take as gifts and were not returnable once taken from the cart for infection control purposes. Funding for the book cart was provided by the Association for Auxiliaries for Children of the Lucile Packard Foundation.

### Clinical and Demographic Measures

To characterize the sample, the following information about the participants was collected from the EMR: Gestational age (weeks), birthweight (g), sex assigned at birth (male = 0, female = 1), length of stay (days), and public insurance. Public insurance was used as a proxy for socioeconomic status (SES), with private insurance indicating higher SES and public insurance indicating lower SES (private = 0, public = 1). In California, eligibility for public insurance is based on a family’s income-to-needs ratio, thus making insurance status an indicator of the economic resources available to a family.^25^

To characterize the health acuity of the sample, we extracted four major comorbidities of prematurity from the EMR: bronchopulmonary dysplasia (BPD), intraventricular hemorrhage (IVH), sepsis, and necrotizing enterocolitis (NEC). BPD was defined as treatment with supplemental oxygen at 36 weeks postmenstrual age (PMA). IVH was defined as the presence of a grade I or higher as per the Papile classification system.^26^ Sepsis was defined as a positive blood culture or >7 days of antibiotics. NEC was defined as a diagnosis of medical or surgical NEC. A binary health acuity score was calculated to group infants into those without these clinical risk factors (health acuity = 0) and infants with one or more of the above comorbidities (health acuity = 1).

### Talk Variables

As part of standard unit practice, bedside nurses documented every instance of family involvement in developmental care activities, including intentional speech directed to the infant, such as reading or talking. By May 2018, standardized protocols guiding developmental care activities and charting procedures were fully operational. These practices were sustained throughout the duration of the current study. Charting of intentional speech activities noted the approximate duration (in minutes) and who participated (e.g., mother, father, staff, or any combination of these). Using these records, we derived six standardized talk metrics that took into account variation in infants’ hospital length of stay. For talk rate, we calculated the total minutes of talk activities divided by the infant’s hospital length of stay (in days). For talk frequency, we calculated the total number of documented talk instances divided by the infant’s hospital length of stay (in days). Each of these measures was generated separately for talk attributed to any family members, talk attributed to only staff, and the combined total from both sources. This yielded six talk metrics in total: total talk rate, family talk rate, staff talk rate, total talk frequency, family talk frequency, and staff talk frequency.

### Data analyses

All analyses were conducted in R version 4.2.2. Descriptive statistics were used to describe all demographic, clinical, and talk-related variables. All variables were assessed for outliers and tested for normality. Values exceeding three standard deviations from the mean were winsorized. Histograms and Shapiro-Wilk tests indicated that family and staff talk rates and frequencies were non-normally distributed; these variables were log-transformed using a base-10 log.

Clinical, socio-demographic and talk metrics were compared between pre- and post-cohorts using chi-square tests for categorical variables and independent samples *t*-tests for continuous variables. To test the effect of the book cart intervention on speech exposure around infants in the NICU, we conducted linear mixed-effects models with repeated observations for each infant, capturing both family and staff talk nested within infants. Health acuity was included as a covariate to account for the possibility that illness severity could influence the overall amount of speech directed to infants. Model 1 included health acuity, talk provider, and book cart presence as fixed effects. Model 2 added a talk provider × book cart interaction term to test whether the effect of the book cart differed for family versus clinical staff members. We performed a secondary sensitivity analysis to assess the robustness of findings related to health acuity. BPD was substituted as a covariate in place of the overall health acuity score, as BPD was the specific diagnosis that differed between groups. Statistical significance for all analyses was defined as *p* < 0.05.

## Results

Table 1 summarizes the demographic, clinical, and talk-related characteristics of NICU infants in the pre- and post-book cart cohorts. No significant clinical or demographic differences were observed, except for BPD, which was more prevalent in the post-book cart cohort. Family and staff in the post-book cart cohort engaged in significantly higher rates and frequencies of talk to the infants compared to those in the pre-book cart group (Table 1).

**Table 1.** Clinical and Demographic Characteristics and Descriptive Statistics of Talking Variables for Pre- and Post-Book Cart Groups

| | Pre-Book cart (n = 66) | | | Post-Book cart (n = 81) | | | *t* or $\chi^2$ |
| --- | --- | --- | --- | --- | --- | --- | --- |
|  | Mean (SD)<br>or n (%) | Min | Max | Mean (SD)<br>or n (%) | Min | Max |  |
| Gestational Age (weeks) | 28.4 (2.3) | 23.1 | 31.8 | 28.5 (2.3) | 23.2 | 31.8 | -0.15 |
| Birthweight (grams) | 1129.9<br>(388.6) | 495.0 | 2463.0 | 1121.6<br>(384.1) | 400.0 | 2224.9 | 0.13 |
| Female | 31 (47.0%) | - | - | 37 (46%) | - | - | 0.00 |
| Length of Stay (days) | 76.7 (38.7) | 24.8 | 196.9 | 73.1 (37.3) | 6.9 | 212.4 | 0.56 |
| Public Insurance<sup>1</sup> | 28 (42.4%) | - | - | 37 (45.7%) | - | - | 0.05 |
| English speaking | 54 (81.8%) | - | - | 68 (84%) | - | - | 0.02 |
| Health Acuity<sup>2</sup> | 32 (48.5%) | - | - | 45 (55.6%) | - | - | 0.47 |
| IVH | 18 (27.3%) | - | - | 17 (21.0%) | - | - | 0.48 |
| BPD | 15 (22.7%) | - | - | 35 (43.2%) | - | - | 5.92* |
| Sepsis | 7 (10.6%) | - | - | 9 (11.1%) | - | - | 0.00 |
| NEC | 17 (25.8%) | - | - | 17 (21.0%) | - | - | 0.23 |
| Talk Rate (minutes/day) | 9.9 (6.3) | 0.0 | 28.7 | 11.9 (5.2) | 0.0 | 27.5 | -2.11* |
| Family Talk Rate | 3.3 (3.5) | 0.0 | 15.5 | 4.6 (3.6) | 0.0 | 15.5 | -2.08* |
| Staff Talk Rate | 6.5 (4.4) | 0.0 | 18.4 | 7.3 (3.2) | 0.0 | 14.7 | -1.19 |
| Talk Frequency (instances/day) | 1.0 (0.5) | 0.0 | 2.2 | 1.2 (0.5) | 0.0 | 2.6 | -2.94** |
| Family Talk Frequency | 0.2 (0.1) | 0.0 | 0.7 | 0.3 (0.2) | 0.0 | 0.8 | -3.18** |
| Staff Talk Frequency | 0.8 (0.4) | 0.0 | 2.0 | 0.9 (0.4) | 0.0 | 2.2 | -2.06* |

<sup>1</sup>Socioeconomic Status (SES) is defined by whether a family has public or private insurance.
<sup>2</sup>Health acuity score is defined by whether the infant has one more of the following conditions: intraventricular hemorrhage (IVH), bronchopulmonary dysplasia (BPD), sepsis, or necrotizing enterocolitis (NEC).
p < 0.10 †; p < 0.05\*; p < 0.01\*\*; p < 0.001\*\*\*

Linear mixed-effects models examined the association between book cart group and talk exposure, adjusting for infant health acuity and talk provider. As shown in Model 1 of Table 2, families and clinical staff engaged in higher rates of talk in the post-book cart group compared to those in the pre-book cart group. Talk provider was also a significant predictor, with clinical staff engaged in talk to infants at higher rates compared to families (Figure 1). Model 2 of Table 2 revealed no significant book cart × talk provider interaction for talk rate.

**Figure 1.**
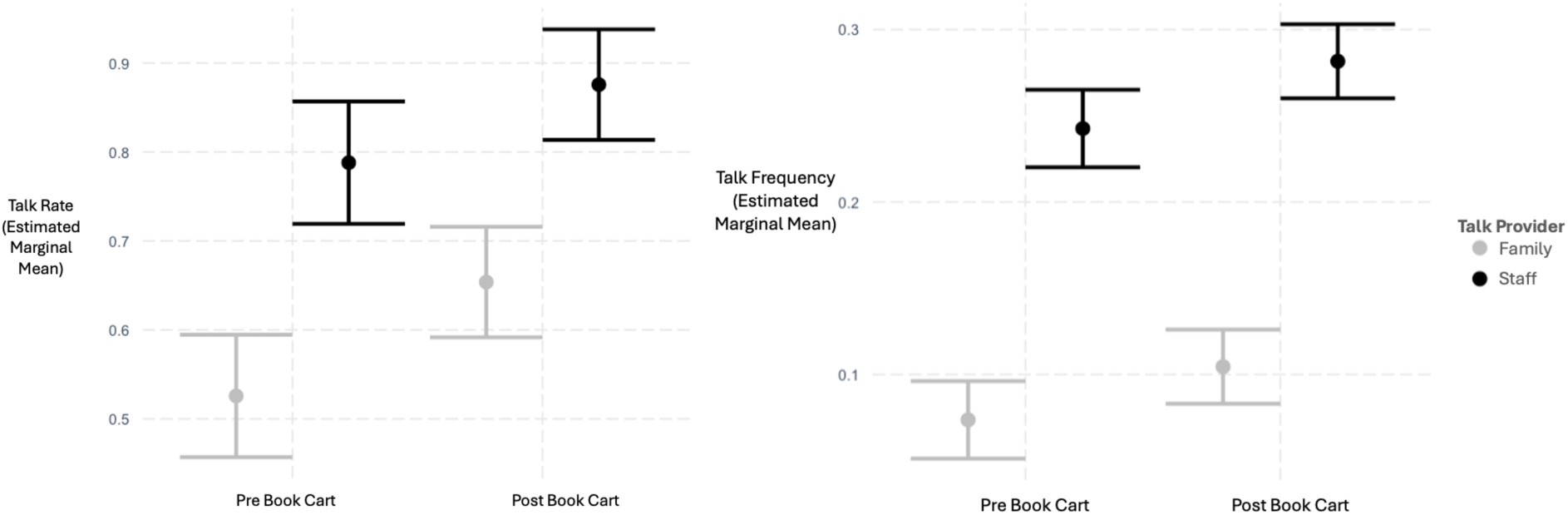
Estimated Marginal Means (95% CI) of Predicted Talking Rates and Frequencies Before and After Book Cart Intervention, by Talk Provider and adjusted for health acuity.

**Table 2.** Linear Mixed Models Predicting Talk Amounts

| Predictor | Talk Rate |  | Talk Frequency |  |
| --- | --- | --- | --- | --- |
|  | Model 1<sup>1</sup><br>β (SE) | Model 2<sup>2</sup><br>β (SE) | Model 3<sup>3</sup><br>β (SE) | Model 4<sup>4</sup><br>β (SE) |
| Health Acuity | 0.06 (0.04) | 0.06 (0.04) | -0.01 (0.01) | -0.01 (0.01) |
| Talk Provider | 0.24 (0.03)*** | 0.26 (0.04)*** | 0.17 (0.01)*** | 0.17 (0.01)*** |
| Book Cart | 0.11 (0.04)** | 0.13 (0.05)** | 0.03 (0.01)** | 0.03 (0.01)* |
| Talk Provider X<br>Book Cart | - | -0.04 (0.05) | - | 0.01 (0.02) |

<sup>1</sup>Main effects model predicting talk rate.
<sup>2</sup>Talk provider X book cart interaction model predicting talk rate.
<sup>3</sup>Main effects model predicting talk frequency.
<sup>4</sup>Talk provider X book cart interaction model predicting talk frequency.
p < 0.10 †; p < 0.05\*; p < 0.01\*\*; p < 0.001\*\*\*

Similarly, as shown in Model 3 of Table 2, there were greater instances of families and clinical staff engaging in talk to infants in the post-book cart group compared to those in the pre-book cart group. Talk provider again predicted talk frequency, with staff engaging in talk to infants more frequently than families (Figure 1). Model 4 of Table 2 showed no significant interaction between book cart presence and talk provider in talk frequency.

Sensitivity analyses re-ran all models, controlling directly for BPD instead of health acuity. Findings were unchanged. Clinical staff and families in the post-book cart group provided significantly higher rates (*β* = 0.11, *SE* = 0.04) and frequencies (*β* = 0.03, *SE* = 0.01) of talk.

Staff talked to infants at higher rates (*β* = 0.24, *SE* = 0.03) and frequencies (*β* = 0.17, *SE* = 0.01) compared to families. No significant talk provider × book cart interactions were found in predicting either talk rate or frequency (all p > 0.05).

## Discussion

This retrospective study found that the introduction of a book cart in the neonatal intensive care unit was associated with higher levels of talk directed to infants born very preterm by both family and staff members. Sensitivity analyses confirmed that findings were not driven by higher incidence of BPD in the post-book cart group. Results of this study suggest that book provision in the NICU may encourage both family members and clinical staff to speak more to preterm infants.

Our findings complement previous experimental studies that have examined the efficacy of reading programs in the NICU, such as Read-a-Thon and read-aloud initiatives. They demonstrate that structured interventions can increase children’s language exposure in the NICU and in the home environment by encouraging families to read to their infants.^17,19,23^ Several nonprofit programs and organizations exist for providing families with books in the NICU.^27–31^ Our study provides evidence that such initiatives can quantifiably enhance the speech environment of the NICU. Facilitating directed speech to infants during NICU hospitalization may set up patterns of behaviors that could persist in the home environment after discharge.

Moreover, providing books in the NICU could be explored as interventions to improve equity in language outcomes, especially for families who may have limited financial resources for purchasing books. Ways to expand the languages of worded books available will help to ensure families from under-represented backgrounds have equitable access to these programs.

Although talk rates and frequencies were higher in the presence of the book cart, overall levels of speech exposure remained low compared to the amount of speech that infants hear in utero. Specifically, caregivers directed between 10 (pre book cart) to 12 minutes (post book cart) of talk per day to infants, rates that are substantially below levels of speech exposure that fetuses of similar gestational ages have been shown to experience in utero^32^. It remains unclear whether the modest difference of 2 minutes per day observed in the current study upon introduction of the book cart would be sufficient to affect infant health or language outcomes. More research is needed to understand the potential mechanisms through which book reading could support brain development and language outcomes, such as through direct enhancement of neural circuit underlying auditory-speech processes and/or through indirect mechanisms that may facilitate caregiver-infant engagement.

Our findings showed evidence that the presence of the book cart was associated with higher engagement in talk directed to infants from both clinical staff and families. This pattern is consistent with our previous work, which shows that clinical staff contribute greater amounts of speech directed to infants than their families, on average ^24^. This finding is not surprising given that clinical staff are regularly present at bedside and involved in all care times, whereas parents face many barriers to being in NICU (i.e., limited parental leave, childcare constraint for other children, travel to hospital) that can limit opportunities to talk to their infant. Supporting language environments of NICU may require multiple strategies, including book carts to encourage live speech or playing recorded speech^33,34^ for periods when families are unable to be present at bedside. Future studies might also test the extent to which including explicit education, instruction, or incentives to parents and staff may enhance the effectiveness of the book cart.^18,22,35^ Providing instructions or educational material alongside reading initiatives can help improve intervention efficacy.^17^ Incentive-based approaches such as progress-tracking with rewards could also encourage more parent engagement in reading in the NICU.^23,35^

This study has limitations. Unlike previous studies,^19,23,36^ which relied on parent-reported reading scores as measures of speech, clinical staff documented amounts of speech in the NICU in-person in real time. The objective nature of our talk measurement confers a significant strength, however, it may introduce inconsistencies in the documentation quality by clinical staff. Awareness of the book cart initiative may have prompted nurses to increase their documentation of talk activities. While the book cart included some wordless picture books, the worded books were only in Spanish and English and the effects were only measured in families who spoke and read in those languages. We were also unable to specifically document whether the talk activities utilized the books from the cart.

## Conclusion

This study shows that family members and staff who had access to a book cart in the NICU talked to infants more frequently and at higher rates than those who did not. The findings illustrated that overall levels of infant-directed talk from both family and staff were low compared to previous estimates of talk experienced in utero, indicating a need to continue to promote opportunities for speech exposure in the NICU. Future studies should evaluate the efficacy of interventions that provide book carts alongside educational information on the importance of reading and in encouraging both families and clinical staff to speak around infants in the NICU. Increased family and staff talk to infants in the NICU may be important for promoting language development and for establishing positive caregiver verbal engagement behaviors.

## Author Contributions

**Sasha Montgomery:** conceptualization, investigation, writing – original draft, methodology, validation, software, formal analysis, data curation, project administration. **Avery Look:** conceptualization, investigation, writing – review and editing, validation, software, formal analysis, data curation, project administration. **Molly Lazarus:** conceptualization, investigation, writing – review and editing, validation, software, formal analysis, data curation, project administration**. Virginia A. Marchman:** conceptualization, investigation, writing – review and editing, validation, methodology, software, supervision, data curation. **Melissa Scala:** conceptualization, investigation, writing – review and editing, methodology, project administration, supervision, resources and funding. **Katherine E. Travis:** conceptualization, investigation, funding acquisition, writing – review and editing, methodology, project administration, data curation, supervision, resources and funding.

## Data Availability

The datasets presented in this article are not readily available because of patient privacy issues but de-identified data are available from the corresponding author on reasonable request and subject to institutional approvals.

## Acknowledgements

We gratefully acknowledge the families who were cared for at our center and the clinical staff whose careful and consistent charting made this work possible. This study was supported by the Association for Auxiliaries for Children of the Lucile Packard Foundation

## Ethics Statement

This study was considered a minimal-risk retrospective chart review; families were not required to provide consent (Stanford University IRB #44480).

## Data Availability Statement

Deidentified individual participant data will be made available upon request.

## References

1. Vandormael C, Schoenhals L, Hüppi PS, Filippa M, Borradori Tolsa C. Language in Preterm Born Children: Atypical Development and Effects of Early Interventions on Neuroplasticity. Neural Plasticity. 2019;2019:1–10. doi:10.1155/2019/6873270

2. Zambrana IM, Vollrath ME, Jacobsson B, Sengpiel V, Ystrom E. Preterm birth and risk for language delays before school entry: A sibling-control study. Dev Psychopathol. 2021;33(1):47–52. doi:10.1017/S0954579419001536

3. Guarini A, Marini A, Savini S, Alessandroni R, Faldella G, Sansavini A. Linguistic features in children born very preterm at preschool age. Develop Med Child Neuro. 2016;58(9):949–956. doi:10.1111/dmcn.13118

4. Van Noort-van Der Spek IL, Franken MCJP, Weisglas-Kuperus N. Language Functions in Preterm-Born Children: A Systematic Review and Meta-analysis. Pediatrics. 2012;129(4):745–754. doi:10.1542/peds.2011-1728

5. McMahon E, Wintermark P, Lahav A. Auditory brain development in premature infants: the importance of early experience. Annals of the New York Academy of Sciences. 2012;1252(1):17–24. doi:10.1111/j.1749-6632.2012.06445.x

6. Ruben RJ. The Ontogeny of Human Hearing. Acta Oto-Laryngologica. 1992;112(2):192–196. doi:10.1080/00016489.1992.11665402

7. Caskey M, Stephens B, Tucker R, Vohr B. Importance of Parent Talk on the Development of Preterm Infant Vocalizations. Pediatrics. 2011;128(5):910–916. doi:10.1542/peds.2011-0609

8. Best K, Bogossian F, New K. Language Exposure of Preterm Infants in the Neonatal Unit: A Systematic Review. Neonatology. 2018;114(3):261–276. doi:10.1159/000489600

9. Karrass J, Braungart-Rieker JM. Effects of shared parent–infant book reading on early language acquisition. Journal of Applied Developmental Psychology. 2005;26(2):133–148. doi:10.1016/j.appdev.2004.12.003

10. Dowdall N, Melendez-Torres GJ, Murray L, Gardner F, Hartford L, Cooper PJ. Shared Picture Book Reading Interventions for Child Language Development: A Systematic Review and Meta-Analysis. Child Development. 2020;91(2). doi:10.1111/cdev.13225

11. Muhinyi A, Rowe ML. Shared reading with preverbal infants and later language development. Journal of Applied Developmental Psychology. 2019;64:101053. doi:10.1016/j.appdev.2019.101053

12. Ece Demir-Lira Ö, Applebaum LR, Goldin-Meadow S, Levine SC. Parents’ early book reading to children: Relation to children’s later language and literacy outcomes controlling for other parent language input. Developmental Science. 2019;22(3):e12764. doi:10.1111/desc.12764

13. Farran LK, Leslie SL, Brasher SN. Promoting language and literacy through shared book reading in the NICU: A scoping review. Dey SK, ed. PLoS ONE. 2025;20(3):e0318690. doi:10.1371/journal.pone.0318690

14. Franks AM, Seaman C, Franks EK, Rollyson W, Davies T. Parental Reading to Infants Improves Language Score: A Rural Family Medicine Intervention. J Am Board Fam Med. 2022;35(6):1156–1162. doi:10.3122/jabfm.2022.220064R2

15. Weitzman CC, Roy L, Walls T, Tomlin R. More Evidence for Reach Out and Read: A Home-Based Study. Pediatrics. 2004;113(5):1248–1253. doi:10.1542/peds.113.5.1248

16. Connor Garbe M, Bond SL, Boulware C, et al. The Effect of Exposure to Reach Out and Read on Shared Reading Behaviors. Academic Pediatrics. 2023;23(8):1598–1604. doi:10.1016/j.acap.2023.06.030

17. King TM, Muzaffar S, George M. The Role of Clinic Culture in Implementation of Primary Care Interventions: The Case of Reach Out and Read. Academic Pediatrics. 2009;9(1):40–46. doi:10.1016/j.acap.2008.10.004

18. Neri E, De Pascalis L, Agostini F, et al. Parental Book-Reading to Preterm Born Infants in NICU: The Effects on Language Development in the First Two Years. IJERPH. 2021;18(21):11361. doi:10.3390/ijerph182111361

19. Lariviere J, Rennick JE. Parent Picture-Book Reading to Infants in the Neonatal Intensive Care Unit as an Intervention Supporting Parent-Infant Interaction and Later Book Reading. Journal of Developmental & Behavioral Pediatrics. 2011;32(2):146–152. doi:10.1097/DBP.0b013e318203e3a1

20. Joaquim P, Calado G, Costa M. Benefits of reading to premature newborns in the neonatal intensive care unit: A scoping review. Journal of Neonatal Nursing. 2024;30(4):325–330. doi:10.1016/j.jnn.2023.11.011

21. Boissel L, Guilé JM, Viaux-Savelon S, et al. A narrative review of the effect of parent–child shared reading in preterm infants. Front Pediatr. 2022;10:860391. doi:10.3389/fped.2022.860391

22. Jain VG, Kessler C, Lacina L, et al. Encouraging Parental Reading for High-Risk Neonatal Intensive Care Unit Infants. The Journal of Pediatrics. 2021;232:95–102. doi:10.1016/j.jpeds.2021.01.003

23. Latif M, Duarte Ribeiro AP, Blatz MA, et al. Encouraging Our NICU to “Read-a-Latte”: Leveraging a Read-a-Thon to Launch a Quality Improvement Initiative. Advances in Neonatal Care. 2023;23(2):120–131. doi:10.1097/ANC.0000000000001038

24. Scala ML, Marchman VA, Godenzi C, Gao C, Travis KE. Assessing speech exposure in the NICU: Implications for speech enrichment for preterm infants. J Perinatol. 2020;40(10):1537–1545. doi:10.1038/s41372-020-0672-7

25. Lazarus MF, Marchman VA, Brignoni-Pérez E, et al. Inpatient Skin-to-skin Care Predicts 12-Month Neurodevelopmental Outcomes in Very Preterm Infants. The Journal of Pediatrics. 2024;274:114190. doi:10.1016/j.jpeds.2024.114190

26. Papile LA, Burstein J, Burstein R, Koffler H. Incidence and evolution of subependymal and intraventricular hemorrhage: A study of infants with birth weights less than 1,500 gm. The Journal of Pediatrics. 1978;92(4):529–534. doi:10.1016/S0022-3476(78)80282-0

27. Babies With Books. https://www.babieswithbooks.org/

28. Little Giraffe Foundation. https://www.littlegiraffefoundation.org/

29. Books For Babies. https://www.booksforbabies.org/

30. Project: Cameron’s Story. https://www.projectcameronsstory.com/

31. Reading Heart. https://www.readingheart.org/

32. Monson BB, Ambrose SE, Gaede C, Rollo D. Language Exposure for Preterm Infants is Reduced Relative to Fetuses. The Journal of Pediatrics. 2023;262:113344. doi:10.1016/j.jpeds.2022.12.042

33. Travis KE, Scala M, Marchman VA, et al. Listening to Mom in the Neonatal Intensive Care Unit: A randomized trial of increased maternal speech exposure on white matter connectivity in infants born preterm. Preprint posted online September 23, 2024. doi:10.1101/2024.09.20.24314094

34. Webb AR, Heller HT, Benson CB, Lahav A. Mother’s voice and heartbeat sounds elicit auditory plasticity in the human brain before full gestation. Proc Natl Acad Sci USA. 2015;112(10):3152–3157. doi:10.1073/pnas.1414924112

35. New chapters unfold for UAB Medicine NICU Bookworms reading program.https://www.uabmedicine.org/news/news/new-chapters-unfold-for-uab-medicine-nicu-bookworms-reading-program/.

36. Fraser A, Griffiths N, Webb A. Why reading matters. The development of a read-a-thon for neonatal intensive care units to encourage neonatal exposure to language. Journal of Neonatal Nursing. 2023;29(5):704–708. doi:10.1016/j.jnn.2023.02.003

